# A descriptive survey of patient experiences and access to specialty medicines with alternative funding programs

**DOI:** 10.1101/2024.06.28.24309668

**Authors:** William B. Wong, Irina Yermilov, Hannah Dalglish, Lori Bienvenu, Jonathan James, Sarah N. Gibbs

## Abstract

**Background:** Alternative funding programs (AFPs) seek to reduce plan sponsor costs by excluding specialty drugs from a beneficiary’s plan coverage and requiring patients to obtain medications through alternative sources (typically, the manufacturer’s patient assistance programs [PAPs]) via an AFP vendor as a third-party).

**Objective:** To describe patients’ experiences and medication access with AFPs, which have not been explored previously.

**Methods:** A survey instrument consisting of optional single- and multiple-choice questions with branching logic was administered to patients recruited from an online patient panel and a patient advocacy group who had experience with AFPs. The survey assessed patients’ awareness of AFPs from their employers, experience with the PAP application process via the AFP vendor, timeliness of medication access (if granted), and/or the health impact of any delay in access. All analyses were descriptive and exploratory subgroup analyses were conducted by disease area and reported income levels.

**Results:** In total, 227 patients were included in the final sample. Most patients (61%) first heard of the AFP as part of their health benefit when trying to obtain their medication. Up to 88% of patients reported being stressed owing to the medication coverage denial and the uncertainty of obtaining their medication. Over half of patients (54%) reported being uncomfortable with the benefits manager from the AFP vendor. On average, patients reported waiting to receive their medication for approximately 2 months (68.2 days); 24% reported the wait for the medication worsened their condition and 64% reported the wait led to stress and/or anxiety. Patients who indicated the wait time negatively affected them had considered a job change or left their job at a 3–5-fold higher rate than those who reported no impact from wait time. Patients with hemophilia and other bleeding disorders reported receiving their prescribed medication less often than patients with other conditions (63% vs 82%), while more patients with lower incomes (< $50,000 vs > $50,000) reported not receiving any medication (12% vs 5%).

**Conclusions:** Most patients who obtain their specialty medicines via AFPs reported being uncomfortable with the process and experiencing treatment delays, which may have been linked to disease progression, worsened mental well-being and consideration of a job change. Employers should be aware of the potential downstream impacts on employee health, retention, and the employee–employer relationship when considering implementing an AFP into their health plan.

**Plain Language Summary:** Patients who have used alternative funding programs (AFPs) to access their medication were surveyed to understand their experiences. We found that using AFPs may lead to delays in patients receiving their medication, which may lead to worsening of their disease and add to their stress/anxiety. Employers should be mindful that, because of AFPs, patients reported considering leaving their jobs to find a role with better insurance coverage.

## Introduction

Specialty medications have traditionally been defined as those that treat chronic, complex, or serious conditions.^1^ While many of these medications improve clinical outcomes, concerns have arisen about their affordability. Consequently, pharmaceutical manufacturers may offer copay assistance to improve affordability and reduce the out-of-pocket cost burden for commercially insured patients.^2^ Alternatively, patient assistance programs (PAPs, free drug programs) or charitable foundations, which can be funded by manufacturers or other private sources, are aimed at supporting patients who are uninsured or underinsured (insured patients with significant financial burden).^3,4^ Although PAPs and charitable foundations generally provide medications free of charge, income restrictions are typically in place, and patients with higher incomes are excluded from these programs.

In recent years, alternative funding programs (AFPs) have emerged as a new way to limit plan sponsors’ exposure (i.e. employers) to the cost of specialty medications. These programs are operated by vendors who work on behalf of plan sponsors to exclude certain specialty medications from a beneficiary’s health plan coverage.^5–7^ The AFP vendors then seek alternative sources to obtain the patient’s medication. Typically, the alternative sources are PAPs or foundations, or they may include sources outside of the United States.^5,7^ The use of AFPs thus far has been limited, with 14% of employers and 7% of health plans reporting currently using AFPs in 2023. However, there is potential for these programs to grow, with an additional 14% of employers and 33% of health plans reporting exploring their use.^8^

A number of concerns have been raised around these programs. There are ethical considerations of diverting limited resources from PAPs and charitable foundations away from patients without insurance, who rely on these programs as a critical safety net and instead give them to insured patients. Furthermore, the AFP process of coverage denial and subsequently applying for aid can take time leading to potential treatment delays and disruption.^4–6^ Lastly, there is additional administrative complexity for patients to obtain their medication via the AFP process, as well as privacy concerns, which may result in a negative experience for plan beneficiaries.^4,9^ Although these concerns are potentially alarming, there has been no systematic research to support these hypotheses to date. To further understand the impact of AFPs, we conducted a patient survey to gather patients’ experiences with the AFP process and their medication access through AFPs.

## Methods

A cross-sectional survey was conducted between October and December 2023. This study used convenience sampling to concurrently recruit participants from the Rare Patient Voice (RPV) patient panels and the Hope for Hemophilia (HOPE) patient advocacy group. In previous studies, RPV patient panels have been used across multiple disease areas,^10–12^ and in the present study they were included to survey patients across conditions that may be treated with specialty medications. The HOPE patient advocacy group was used primarily to survey patients with hemophilia, because there have been anecdotes of these patients being impacted by AFPs.^13,14^ RPV used a panel method to prevent duplicate responses, and duplicate responses from HOPE were mitigated via Internet Protocol (IP) tracking from Qualtrics, which prevented respondents from the same IP address completing the survey twice. Additionally, patient demographic responses were evaluated for potential duplicative participation from each data source. Respondents received compensation for their participation.

To identify patients who had experience with AFPs, we developed a 4-item screening tool (Supplementary Table 1). Patients were required to have employer- or union-sponsored health insurance and a chronic condition requiring a specialty medication. The specialty medication had to be excluded from their insurance coverage (but not if it was part of step therapy), and patients had to acquire it by contacting an AFP vendor to help them to enroll in a PAP. Only adults (aged > 18 years) were eligible to complete the survey, including caregivers of patients aged < 18 years. Eligible patients were then invited to complete a 26-item survey comprising single- and multiple-choice questions, any of which patients could opt out of answering. Although the survey was not formally pilot tested, the content was reviewed by HOPE for comprehension from a patient perspective. The study protocol, screening tool, and survey were reviewed and approved by the Western Institutional Review Board.

The survey was administered via Qualtrics, and data were analyzed descriptively (proportions, means and medians) using SAS version 9.4 (SAS Institute Inc); no statistical analyses were conducted. Where a participant skipped optional questions, this was considered missed data and excluded. Exploratory subgroup analyses were conducted by disease area (for those subgroups with ≥ 30 respondents) and income (< $50,000 vs > $50,000).

## Results

### PATIENT DEMOGRAPHICS

In total, 7,546 patients were screened, of whom 6,828 were recruited from RPV and 718 from HOPE. Of these, 227 patients had experience with AFPs, provided consent, and answered at least one question in the survey (Supplementary Table 1). Most patients were 18 years or older (90%), male (70%), non-Hispanic white (71%), and lived in a suburb near a large city (43%) (Table 1). The most common health conditions reported were multiple sclerosis (22%), cancer (15%), and hemophilia/bleeding disorders (14%). Around a quarter of patients (27%) reported annual income < $50,000, 61% > $50,000, and 12% did not wish to report or did not know their income.

**Table 1.** Survey Sample Demographics.

| **Patient characteristics** | **n (%)<sup>a</sup>** |
| --- | --- |
| **Total** | 227 (100) |
| **Age, years** | 211 |
| < 18 | 21 (10) |
| 18–34 | 61 (28.9) |
| 35–44 | 46 (21.8) |
| 45–54 | 50 (23.7) |
| ≥ 55 | 32 (15.2) |
| Do not wish to report | 1 (0.5) |
| Unknown | 16 |
| **Gender** | 207 |
| Female | 61 (29.5) |
| Male | 144 (69.6) |
| Do not wish to report | 2 (1.0) |
| Unknown | 20 |
| **Race and ethnicity, n** | 211 |
| Asian/Pacific Islander/American Indian or Alaska Native<sup>b</sup> | 5 (2.4) |
| Black<sup>b</sup> | 18 (8.5) |
| Hispanic, Latino or Spanish origin of any race | 22 (10.4) |
| Race/ethnicity not listed or do not wish to report | 12 (5.7) |
| Two or more races<sup>b</sup> | 4 (1.9) |
| White<sup>b</sup> | 150 (71.1) |
| Unknown | 11 |
| **Yearly income, n** | 211 |
| < \$25,000 | 19 (9.0) |
| \$25,000–\$50,000 | 38 (18.0) |
| \$50,000–\$75,000 | 44 (20.9) |
| \$75,000–\$100,000 | 46 (21.8) |
| > \$100,000 | 39 (18.5) |
| Do not wish to report or do not know | 25 (11.8) |
| Unknown | 16 |
| **Type of community, n** | **209** |
| Large city | 46 (22.0) |
| Suburb near a large city | 89 (42.6) |
| Small city or town | 49 (23.4) |
| Rural area | 24 (11.5) |
| Do not wish to report | 1 (0.5) |
| Unknown | 18 |
| **Health condition, n<sup>c</sup>** | **211** |
| Arthritis | 21 (10.0) |
| Cancer | 32 (15.2) |
| Crohn's disease, ulcerative colitis, or other GI disease | 18 (8.5) |
| Hemophilia, or other bleeding disorder | 30 (14.2) |
| Multiple sclerosis | 47 (22.3) |
| Skin condition (such as psoriasis or eczema) | 10 (4.7) |
| Other rare disease not mentioned above | 38 (18.0) |
| Other non-rare disease not mentioned above | 9 (4.3) |
| Do not wish to report | 6 (2.8) |
| Unknown | 16 |

*'Unknown' are respondents who did not answer the question.*
*<sup>a</sup>Proportions may not total 100 owing to rounding.*
*<sup>b</sup>Not Hispanic, Latino, or Spanish origin.*
*<sup>c</sup>Condition that a patient's excluded specialty medication was intended to treat.*
*GI = gastrointestinal.*

### PATIENT AWARENESS OF AFPS AS PART OF HEALTH INSURANCE COVERAGE

Most patients (61% [136/223]) reported that they first learned about AFPs when they attempted to obtain their specialty medication and discovered it was excluded from their health plan (Figure 1). Overall, 28% (62/223) of patients reported being told about AFPs from their employer, including 19% (42/223) of patients who reported their employer let them know an AFP would automatically be applied to all their employees’ health plan, or were strongly encouraged or forced to enroll in the AFP. Among patients encouraged or forced to enroll in AFPs, over half (51% [20/39]) reported being uncomfortable with the pressure from their employer (Figure 2). Furthermore, more than half of patients (54% [115/213]) were uncomfortable discussing their medication needs or financial challenges accessing their medication with their employer.

**Figure 1.**
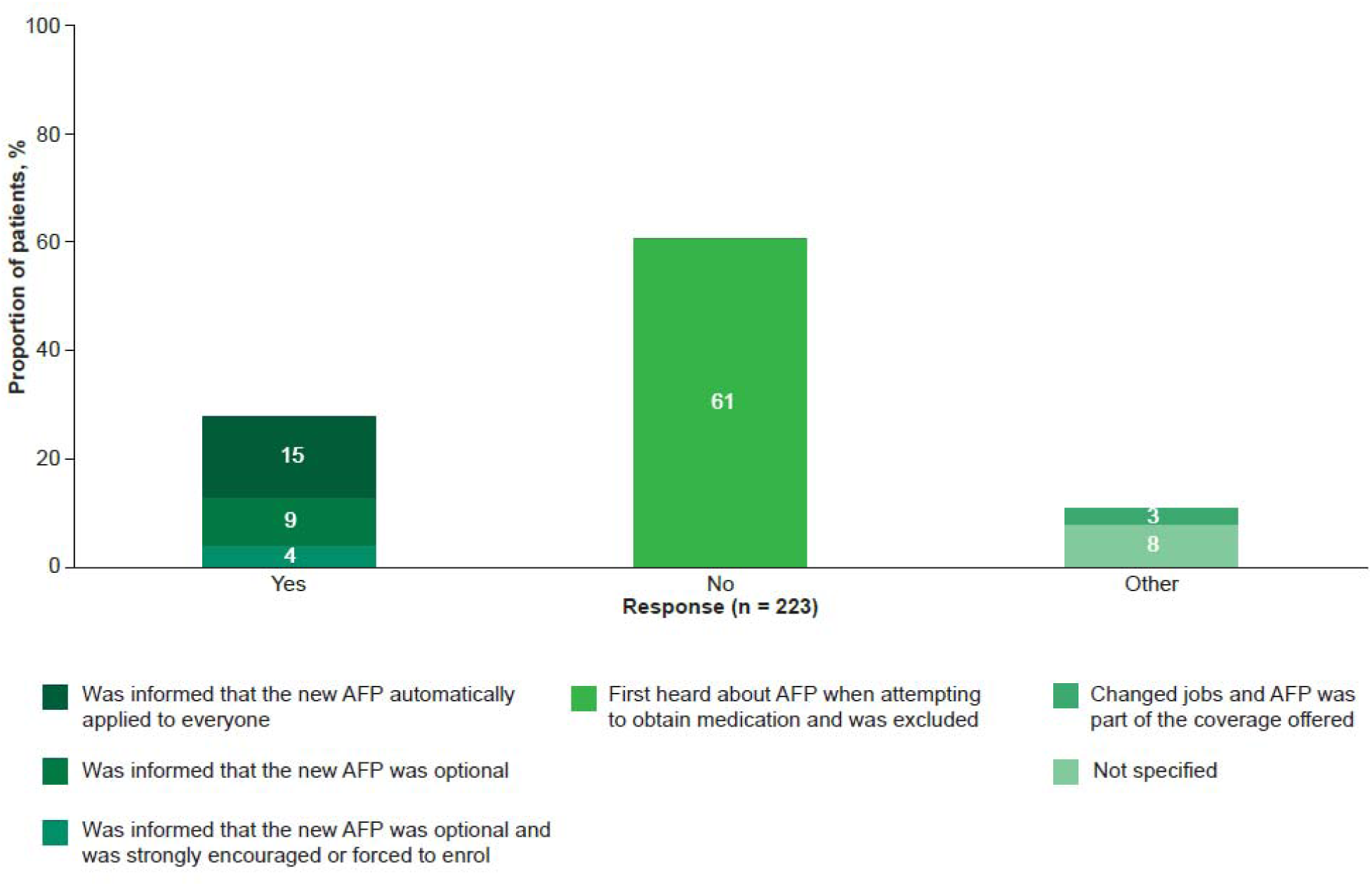
Patient Awareness of AFP Program Which Would Impact their Specialty Medication Coverage. AFP = alternative funding program.

**Figure 2.**
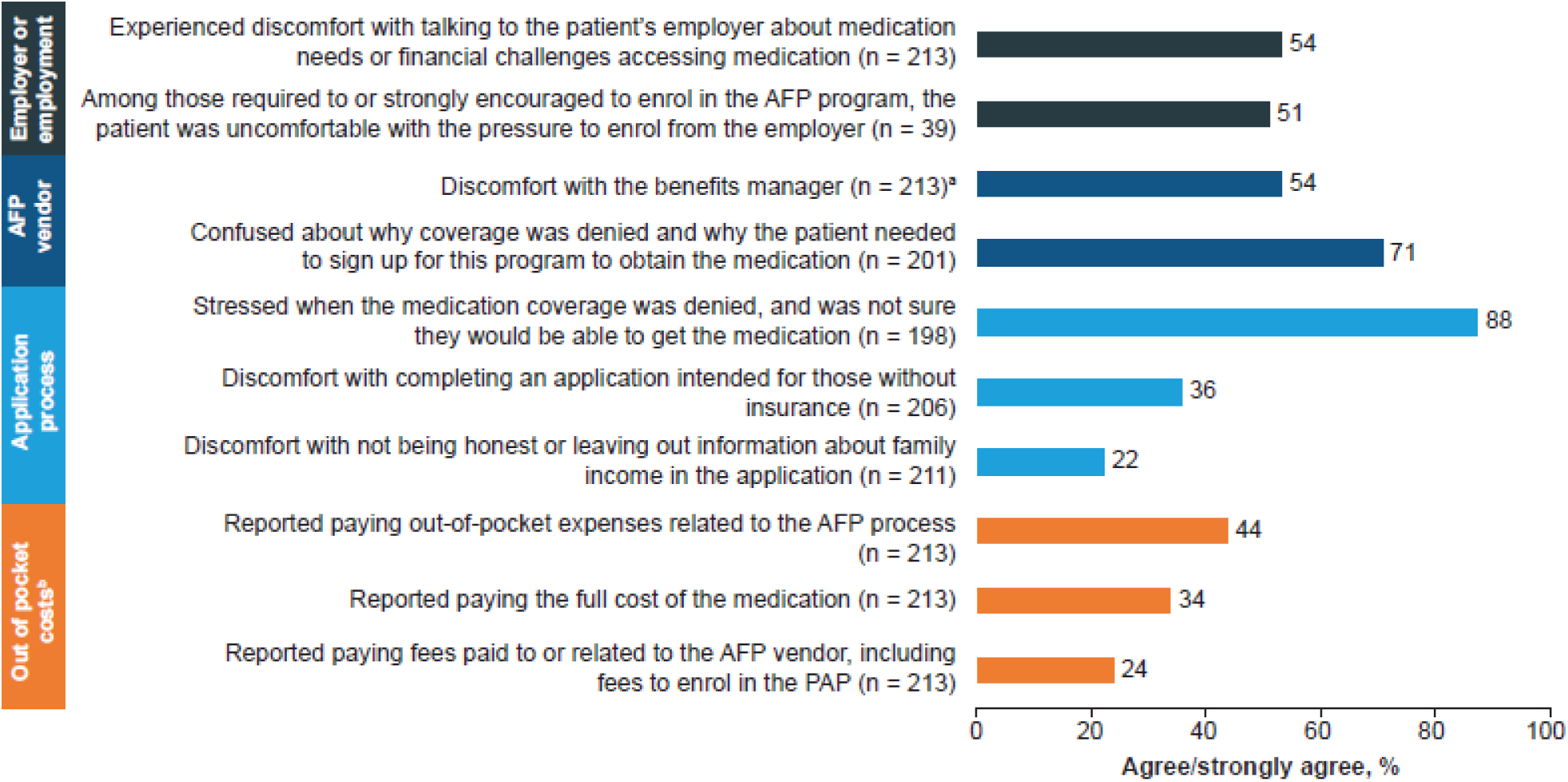
Patient Experiences with Employer, AFP Vendor, and PAP Application Process. ^a^Talking to them about medication needs or financial challenges, providing them with sensitive information, or confused about who they were. ^b^These statements were multiple choice, and all that were true could be selected. AFP = alternative funding program; PAP = patient assistance programs.

### PATIENT EXPERIENCES WITH THE AFP VENDOR AND PAP APPLICATION PROCESS

Almost 9 out of 10 patients (88% [174/198]) reported being stressed by their medication coverage being denied and the uncertainty of obtaining their medication. Additionally, 71% (143/201]) of patients reported confusion over why their coverage was denied and why they needed to sign up with the AFP vendor to obtain their medication. Over half of patients (54% [115/213]) reported being uncomfortable with the benefits manager from the AFP vendor for one or more reasons, including medication needs (26% [30/115), financial challenges (27% [31/115]), providing sensitive information (31% [36/115]), and confusion as to who they were (40% [46/115]). Lastly, 44% (94/213) of patients reported paying an out-of-pocket expense related to the AFP process, including 34% (72/213) who paid the full cost of the medication and 24% (51/213) who paid fees related to the AFP vendor (including fees to enroll in the PAP).

### PATIENTS’ ACCESS TO SPECIALTY MEDICATION

On average, patients reported a mean waiting time to receive their medication of approximately 2 months or 68.2 days and a median of 1.5 months. Patients indicated that the delay in receiving medication had negative impacts, with 24% (51/215) reporting that their condition worsened and 64% (138/215) reporting that the wait led to stress and/or anxiety (Table 2). The mean wait time was approximately 2 times longer for patients with worsened condition or stress and/or anxiety resulting from wait time than patients who reported no impact (95.3 and 71.3 days vs 43.0 days, respectively). These patients also reported considering a job change or leaving their job at 3–5-fold higher rates than those who reported no impact from the wait time (considered leaving job or left their job owing to health insurance, respectively: worsened condition, 38% [18/48] and 20% [9/46]; stress and/or anxiety, 34% [44/128] and 13% [16/128]; no impact, 7% [3/43] and 4% [2/48]).

**Table 2.** Impact of Waiting for Specialty Medication.

| Measure | Overall | Impact of wait for specialty medication |  |  |
| --- | --- | --- | --- | --- |
|  |  | Worsened condition | Stressed/anxious | No impact |
| **Impact of wait for specialty medication, n (%)** | 215 (100) | 51 (24) | 138 (64) | 49 (23) |
| **Time to receiving or waiting for medication, n** | 200 | 48 | 129 | 44 |
| Mean ± SD, days | 68.2 ± 72.7 | 95.3 ± 96.2 | 71.3 ± 76.5 | 43.0 ± 41.7 |
| Ratio of worsened condition or stressed/anxiety to no impact | – | 2.2 | 1.7 | – |
| **Considered leaving their job owing to health insurance, n** | 198 | 48 | 128 | 43 |
| n (%) | 59 (29) | 18 (38) | 44 (34) | 3 (7) |
| Ratio of worsened condition or stressed/anxiety to no impact | – | 5.4 | 4.9 | – |
| **Left their job due to health insurance** | 202 | 46 | 128 | 48 |
| n (%) | 26 (13) | 9 (20) | 16 (13) | 2 (4) |
| Ratio of worsened condition or stressed/anxiety to no impact | – | 4.7 | 3.0 | – |

*Proportions are based on respondents who answered whether the wait for their medication had an impact on their health and the subsequent question of interest in the table rows. SD = standard deviation.*

### EXPLORATORY ANALYSES BY DISEASE AREA AND INCOME

Compared with all other respondents, a lower proportion of patients with hemophilia reported receiving their originally prescribed medication (82% vs 63%, respectively) and having their initial PAP application approved (67% vs 26%) (Table 3). Additionally, compared with all other patients, a greater proportion of patients with hemophilia reported being stressed and/or anxious as a result of waiting for their medication (60% vs 90%, respectively), not receiving any medication (4% vs 23%), and reported longer mean waiting times to receive their medication (66.0 vs 83.7 days, respectively).

**Table 3.**
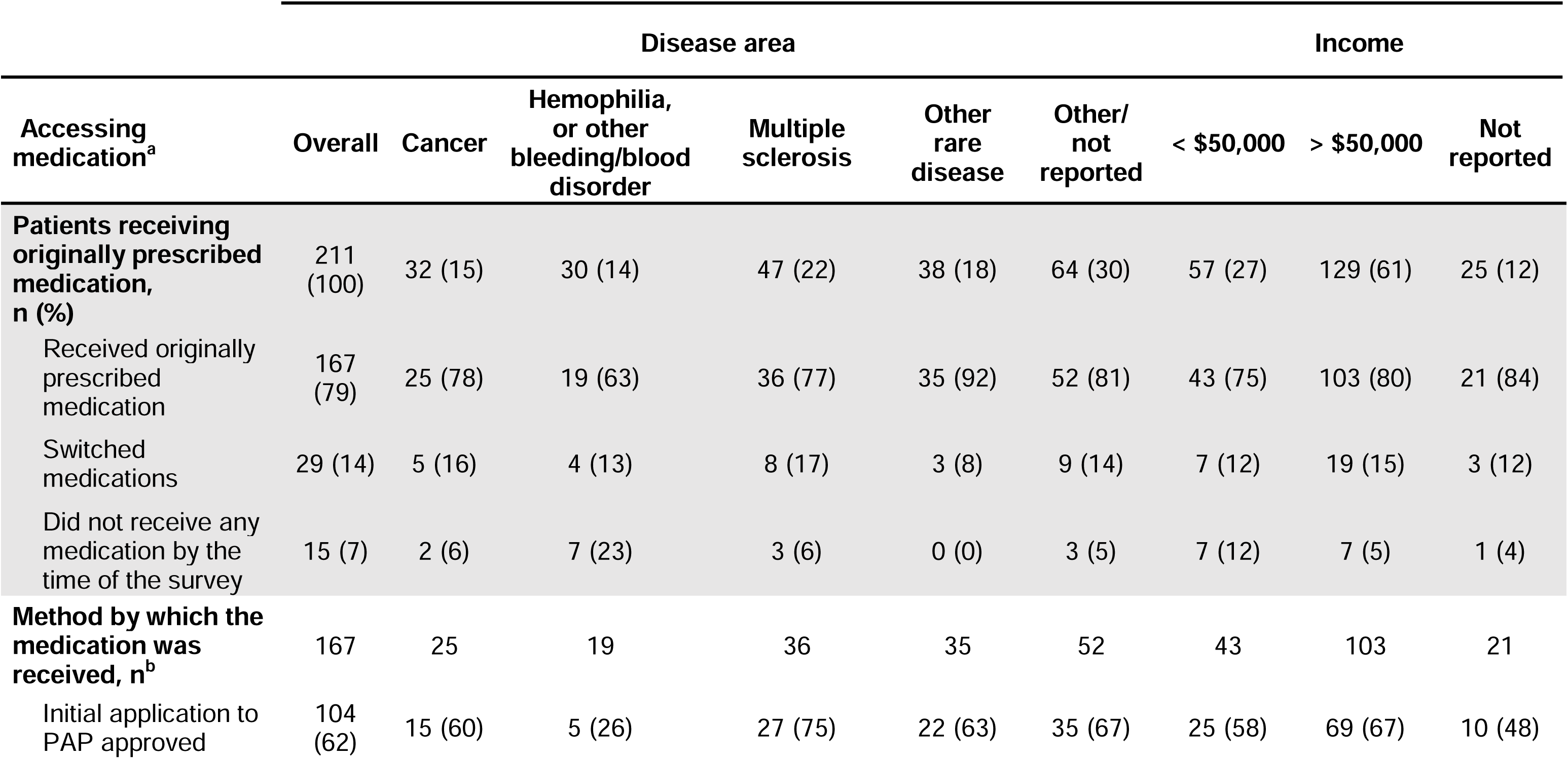
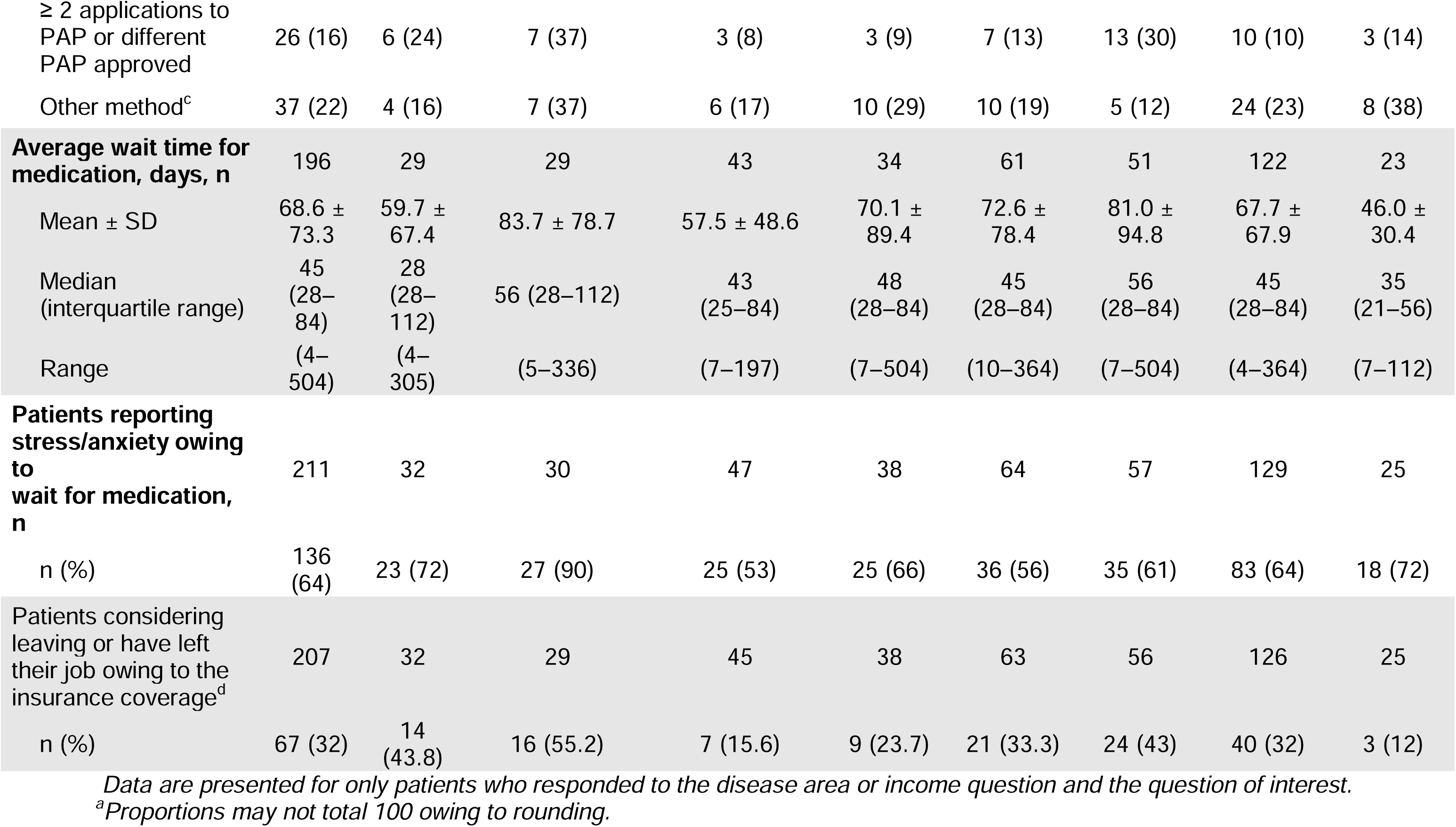
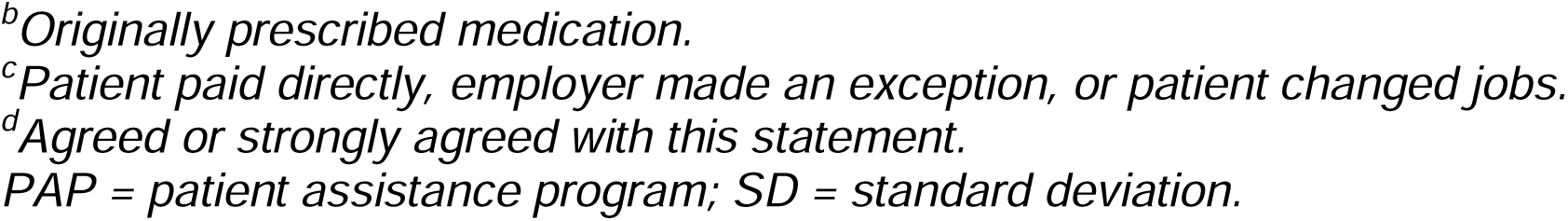
Exploratory Analyses by Disease Area and Income Levels.

Compared with patients reporting an income > $50,000, a greater proportion of patients with incomes < $50,000 reported not receiving their medication at all (5% vs 12%) (Table 3). Furthermore, patients with lower incomes waited longer mean times for their medication than patients with higher incomes (81.0 vs 67.7 days) and reported considering leaving or having left their jobs owing to their insurance coverage at a higher rate (43% vs 32%).

## Discussion

In this cross-sectional descriptive survey, we found that the AFP process added confusion and complexity for patients seeking to obtain their medication. Some patients waited a long time to obtain their medicine, which may have caused them additional stress and worsened their health conditions. To our knowledge, this is the first study examining patients’ experience with and the impact of AFPs, including their impact on access to specialty medications. These findings have implications for both employers and their employees.

Our findings detailing the delays in patients accessing their medication aligns with previous commentaries that have hypothesized that AFPs might result in treatment delays and/or disruption.^4,5^ We found that the average wait time for patients to receive their medication was approximately 2 months (median 1.5 months), which is considerably longer than the wait time reported in the literature to obtain cancer medications without AFP involvement (median 6–15 days)^15,16^ or specialty medications within specialty pharmacies (means of 2–7 days).^17–19^ Given the seriousness of the conditions treated by specialty medications, delays in accessing medication may have significant clinical consequences. In metastatic non-small cell lung cancer, previous research has shown that a delay in treatment initiation of as little as 3 weeks may be associated with a > 2-fold higher risk of death.^20^ In early stage cancers, delays in adjuvant treatment may be associated with up to a 13% higher risk of death.^21^ Overall, 24% of respondents within our survey self-reported that their condition worsened as a result of waiting for their medication. Additionally, it should be noted that across all conditions reported in this study, most patients reported greater stress and/or anxiety, and many patients with chronic illnesses already have or develop mental health conditions as a result of their disease.^22^ Therefore, close attention should be paid to supporting the mental health of patients using AFPs to access their specialty medicine.

In exploratory subgroup analyses, we found trends suggesting that patients’ experiences may vary by disease state. In particular, patients with hemophilia may experience more challenges accessing their medicine, longer delays, and heightened stress and/or anxiety. Delays or interruption in hemophilia treatment are impactful because regular treatment prophylaxis is associated with lower bleed rates compared with on-demand treatment.^23^ Furthermore, compared with the general population, patients with hemophilia have been shown to have an increased risk of mental health conditions such as depression and anxiety.^24^ Additional stress and/or anxiety among patients with hemophilia may worsen quality of life and be associated with a greater likelihood of bleeds and hospital visits.^25^

In addition to the need for employers and plan sponsors to support their beneficiaries’ or employees’ mental health, our study findings have several other implications. First, most patients reported a lack of awareness regarding the change in their health plan in requiring them to use an AFP vendor to obtain their medication. This suggests that there is a continued need for employers to be more mindful about sharing these updates with their employees. Furthermore, patients reported being uncomfortable with several topics related to the AFP process, including discussing personal information (such as health or finances) with their employer, feeling pressure to enroll in the AFP, and the AFP vendor themselves. Taken together, these findings suggest that AFPs may negatively impact the employee–employer relationship. This is further supported by the proportion of patients who considered leaving or actually left their job, especially among those whose condition worsened or who reported stress and/or anxiety due to the wait for their medication. This may have particularly important implications in job markets in which competition for talent and employee retention is critical. Lastly, stratified by income, our results provide a potential signal that higher and lower wage employees may have different experiences with obtaining their medications via AFP vendors. Although these findings are exploratory and should be interpreted with caution, it may be important for employers to consider whether the addition of AFPs into their health benefits could lead to disparities in access to specialty medication for their employees. Additional research is warranted to further explore any potential discriminatory effect AFPs may have for patients.

## Limitations

The survey methodology used in this study has a number of limitations to consider, including being self-reported (therefore prone to bias), using a convenience sample from two different sources, and limited sample size. The inclusion of a control group to enable comparisons and understand potential biases would have been ideal; however, this was not possible in this study in part owing to the lack of prior information available on potential sample size, because there were no previous studies at the time with a similar design. Direct comparisons were not possible in this study, but for some metrics (such as time to obtaining treatment), we were able to reference the literature to contextualize our results. Nonetheless, future research examining the impact of AFPs should consider the inclusion of a control group to better understand differences in delays in medication access and its effects. In addition, our study was limited in sample size owing to the relatively low prevalence of AFPs, despite attempting to maximize the sample size by screening over 7,500 patients from two data sources. Additionally, the branching and optional questions led to smaller response numbers for certain questions. Since subgroup analyses were limited in sample size, with only 30 patients with hemophilia participating, these should be considered exploratory. Despite the sample size limitations, this study offers the first insights into patients’ experiences with AFPs and can therefore be considered foundational for other research to leverage and expand upon. Lastly, given that a convenience sample of self-reporting patients was used, the generalizability of the results may be limited and only be applicable to those who answered the survey. Because this is the first study surveying patients who had experience with AFPs, the direction of any potential bias is unknown. Further research with additional populations is needed to be able to compare these findings.

## Conclusions

Most patients who obtain their specialty medicines via AFPs reported being uncomfortable with the process and experienced treatment delays, which may lead to disease progression, additional stress and/or anxiety, and consideration of a job change. Employers should be aware of the potential downstream impacts on employee retention and the employee–employer relationship when considering implementing an AFP into their health plan.

## Data Availability

All data produced in the present study are available upon reasonable request to the authors

## Acknowledgments

Medical writing support was provided by Rebecca Spencer Martín, MSci, and Rebecca Hornby, PhD, of Oxford PharmaGenesis, Oxford, UK, with funding from Genentech, Inc.

## Notes

### Competing Interest Statement

William B. Wong is an employee of Genentech, Inc. and has Roche stock/stock options. Irina Yermilov, Hannah Dalglish, and Sarah N. Gibbs are employees of PHAR (Partnership for Health Analytic Research), which was paid by Genentech, Inc. to conduct the research described in this manuscript.

### Funding Statement

This study was funded by Genentech, Inc. Genentech, Inc. were involved in conducting the study.

### Author Declarations

The study protocol, screening tool, and survey were reviewed and approved by the Western Institutional Review Board.

