## Supplementary Table 1. for "A descriptive survey of patient experiences and access to specialty medicines with alternative funding programs"

### Supplementary Appendix

#### Supplementary Table 1. Screener Attrition Table

| Screener Attrition Steps | n (% of previous step) |
|---|---|
| Total patients screened[a] | 7,546 |
| Employer- or union-sponsored insurance | 3,686 (48) |
| Chronic condition treated with specialty medication | 3,230 (88) |
| Specialty medication excluded from coverage[b] | 365 (11) |
| Contact with AFP vendor to help enroll in free drug program | 231 (63) |
| Provided consent and answered ≥ 1 question | 227 (98) |

[a]*HOPE, n = 718; RPV, n = 6,828.*
[b]*Not if it was part of step therapy.*
*AFP = alternative funding program; HOPE = Hope for Hemophilia; RPV = Rare Patient Voice.*
